# Deep Learning-based Phylodynamic Inference of Oropouche Virus Transmission Dynamics in Brazil Between 2023 and 2025

**DOI:** 10.64898/2026.11.24.26368826

**Authors:** Ighor Arantes, Gonzalo Bello, Felipe Gomes Naveca, Edson Delatorre, Antonio Silva Lima Neto, Sebastian Duchene, Anna Zhukova

## Abstract

Oropouche virus caused an unprecedented epidemic across the Americas during 2023–2025, associated with the expansion of the reassortant M_1_L_2_S_2_ lineage. To investigate its dissemination, we develop the Birth–Death Exposed–Infectious Vector–Human (BDEIVH) model within a deep-learning framework that explicitly represents alternating human–vector transmission and incubation periods. The model defines two directional reproduction numbers, R_VH_ and R_HV_, whose product, R_HH_, quantifies the expected number of secondary human infections generated by a single infectious human through one complete transmission cycle. In simulations, BDEIVH provides more accurate R_HH_ estimates than conventional birth–death models, with predictive accuracy improving with phylogeny size and remaining robust under reduced phylogenetic resolution. Across three major Brazilian OROV genomic clusters, estimated durations of infection range from 15.4 to 18.5 days in vectors and from 9.42 to 12.3 days in humans, whereas whole-cluster R_HH_ estimates range from 1.15 to 1.81. R_HH_ estimates reached a peak of 4.4 in the endemic Amazon region and 3.0 in the non-endemic Atlantic Forest region. These findings show that explicitly modelling vector-mediated transmission improves phylodynamic inference and reveals substantial OROV transmission in both endemic and non-endemic regions.

## 1 Introduction

Oropouche virus (*Orthobunyavirus oropoucheense*; OROV) is an arthropod-transmitted virus first isolated in 1955 in Vega de Oropouche, Trinidad and Tobago [1, 2]. OROV is an enveloped, negative-sense RNA virus with a tripartite genome comprising large (L), medium (M), and small (S) segments [3]. It is the aetiological agent of Oropouche fever (OROF), an acute febrile illness characterized primarily by fever, headache, and retro-orbital pain [4]. On the basis of its arthropod-mediated spread, OROV is ecologically classified as an arbovirus [1] (a portmanteau of *ar*thropod-*bo*rne *virus*), a designation encompassing viruses maintained in nature through alternating infections of vertebrates and arthropod vectors, with viral replication occurring in both hosts [5, 6]. OROV circulates through distinct but interconnected sylvatic and urban cycles [1]. In the sylvatic cycle, potential vertebrate hosts include non-human primates, sloths, and birds [1, 7–9], while several species of haematophagous arthropods have been implicated as vectors [1, 7]. Although humans are not required for the long-term maintenance of OROV in nature, spillover events can introduce the virus into human populations, where sustained transmission may become established [7]. In this cycle, OROV is primarily associated with the biting midge *Culicoides paraensis*, the main vector mediating spread among humans [10–12].

From 1955 through 2021, OROF was reported in more than 10 countries and territories, with an estimated 500,000 cases and over 30 documented outbreaks, predominantly in the northern Brazilian states of Amazonas, Pará, and Amapá [13]. Between 2023 and 2025, an outbreak of OROF of unprecedented geographic reach resulted in over 55,000 reported cases across the Americas [14], more than 25,000 of which were reported in Brazil [15]. This outbreak was associated with the expansion of a novel reassortant lineage, designated *M*_1_*L*_2_*S*_2_, which is estimated to have emerged in the northern Brazilian state of Amazonas between 2013 and 2014 [16]. This lineage subsequently spread throughout northern Brazil [16–18] and into other parts of the country where autochthonous OROV transmission had not previously been documented [19–26]. The geographic range of the outbreak further expanded into other countries in South America [27–30] and Central America [31–33], while travel-associated cases were reported in North America [34, 35] and Europe [36–38]. The outbreak was also characterized by the first reported deaths attributed to OROF [39] and the first evidence of OROV vertical transmission [40–42]. Available evidence suggests that the *M*_1_*L*_2_*S*_2_ lineage does not exhibit greater viral fitness [43] or distinct pathogenesis [44] than OROV lineages previously circulating in South America. These findings support the hypothesis that ecological factors, rather than intrinsic changes in viral phenotype, played a major role in driving the geographic expansion and magnitude of the outbreak [25, 45].

Despite the characterization of the spatiotemporal spread and clinical features of the *M*_1_*L*_2_*S*_2_ lineage, key epidemiological aspects of its dissemination remain poorly understood, particularly potential variation in transmissibility across geographic areas. Phylodynamics provides a methodological framework for addressing such questions by integrating phylogenetic patterns with mathematical models of pathogen transmission, thereby enabling the characterization of individual transmission clusters [46, 47]. Inference within this framework has been developed around two modeling approaches: coalescent–based models [48–50] and birth–death (BD) models [51–53], with the latter explicitly describing the forward-time processes of transmission, removal, and sampling that give rise to pathogen phylogenies. Since its original formulation for directly transmitted pathogens [52], BD models have been extended to accommodate more complex dynamics, including incubation periods [53–55] and superspreading events [54, 56]. BD models have also previously been applied to the transmission of arboviruses, however, these studies relied on models that did not explicitly represent the alternating vertebrate–arthropod–vertebrate transmission processes that underlie arboviral spread [57, 58]. Oversimplifying pathogen transmission in phylodynamic models has the potential to bias epidemiological parameter estimates, underestimating pathogen transmissibility [59–62].

From a biological standpoint, a comprehensive phylodynamic model for an arbovirus such as OROV would explicitly represent the host–vector transmission cycle within a multi-type birth–death (MTBD) framework, with infected vertebrate hosts and vectors involved in sylvatic and urban transmission represented as distinct types linked by directional host-to-vector and vector-to-host transmission processes. Implementing such a model would face substantial practical challenges, particularly the near absence of genomic sampling from components of the OROV sylvatic cycle. Moreover, explicitly representing the full host–vector system would increase the computational burden of likelihood-based inference by expanding the system of coupled equations required to evaluate the likelihood [55, 63]. Such computational challenges have motivated the development of likelihood-free, simulation-based approaches to inference. These methods rely on the simulation of data under a model without requiring explicit evaluation of its likelihood, and include Approximate Bayesian Computation (ABC) [64–67] and Deep Learning (DL) [56, 68].

The pioneering application of DL to phylodynamics was developed by Voznica *et al.* through PhyloDeep [56], a supervised, simulation-based framework that used dated pathogen phylo-genies represented by summary statistics or compact bijective ladderized vectors to perform epidemiological parameter estimation. Building on this approach, *Perez and Gascuel* introduced PhyloCNN [68] that represents phylogenies through the local neighborhoods of their internal nodes and tips and employs a convolutional neural network architecture for parameter estimation, achieving comparable or greater accuracy while requiring one to two orders of magnitude fewer simulated training trees than previous DL-based methods. To date, DL-based phylodynamic approaches have been used to model the transmission of SARS-CoV-2 [61, 69, 70], Influenza virus [70] and HIV-1 [56, 61, 68].

Here, we develop a phylodynamic framework tailored to arboviruses and apply it to investigate the dissemination of the OROV *M*_1_*L*_2_*S*_2_ lineage through four objectives: (*i*) to develop a phylodynamic model informed by arboviral transmission dynamics and parameterized using OROV-specific data; (*ii*) to implement this parameter-rich model within a deep-learning framework and assess the identifiability of its parameters; (*iii*) to compare its performance with that of established phylodynamic models; and (*iv*) to apply the resulting framework to OROV genomic clusters circulating in Brazil.

## 2 Results

### 2.1 New model accounting for vector-human transmission (BDEIVH)

To enable the inference of epidemiological parameters from OROV genomic clusters, we developed a phylodynamic model explicitly designed to capture the vector-mediated transmission dynamics of arboviruses (Fig. 1a). The Birth–Death Exposed–Infectious Vector–Human (BDEIVH) model (Fig. 1b) is an MTBD model [53] comprising four epidemiological states: exposed and infectious vectors (*V_E_* and *V_I_*, respectively) and exposed and infectious humans (*H_E_* and *H_I_*, respectively). Within each host type, exposed individuals progress to the infectious state at transition rates *μ_V_* and *μ_H_*, whereas vectors and humans are linked by directional transmission rates *λ_VH_* and *λ_HV_*. The model is parameterized by two directional reproduction numbers, *R_VH_* and *R_HV_*, representing vector-to-human and human-to-vector transmission. Their product defines the composite human-to-human reproduction number, *R_HH_* (Fig. 1c), which represents the expected number of secondary human infections generated through one complete vector-mediated transmission cycle. For parameter inference, we implemented the BDEIVH estimator within a neural-network framework adapted from PhyloCNN [68]. Using parameter ranges tailored to OROV transmission (Table 1), we simulated a total of 412,000 trees under the BDEIVH model (Fig. S1a–j). These were partitioned into independent datasets used to (*i*) train (*n* = 400,000); (*ii*) calibrate the 95% prediction intervals (PIs) (*n* = 8,000); and (*iii*) evaluate the predictive performance of the DL estimator (*n* = 4,000). Within each dataset, trees were further stratified into four tree-size categories (30–50, 50–150, 150–250, and 250–350 tips) with equal numbers of trees assigned to each category. To approximate the limited phylogenetic resolution of empirical OROV genomic data, we generated an OROV-like counterpart of the transmission trees dataset by collapsing branches with insufficient genetic divergence. The DL estimator predicts six epidemiological quantities: the directional reproduction numbers *R_VH_* and *R_HV_*; the composite human-to-human reproduction number *R_HH_*; the total durations of infection in vectors (*D_V_*) and humans (*D_H_*), each encompassing an incubation period and an infectious period; and the vector-to-human ratio (*V*/*H*). Each estimate is accompanied by a 95% prediction interval (PI).

**Figure 1:**
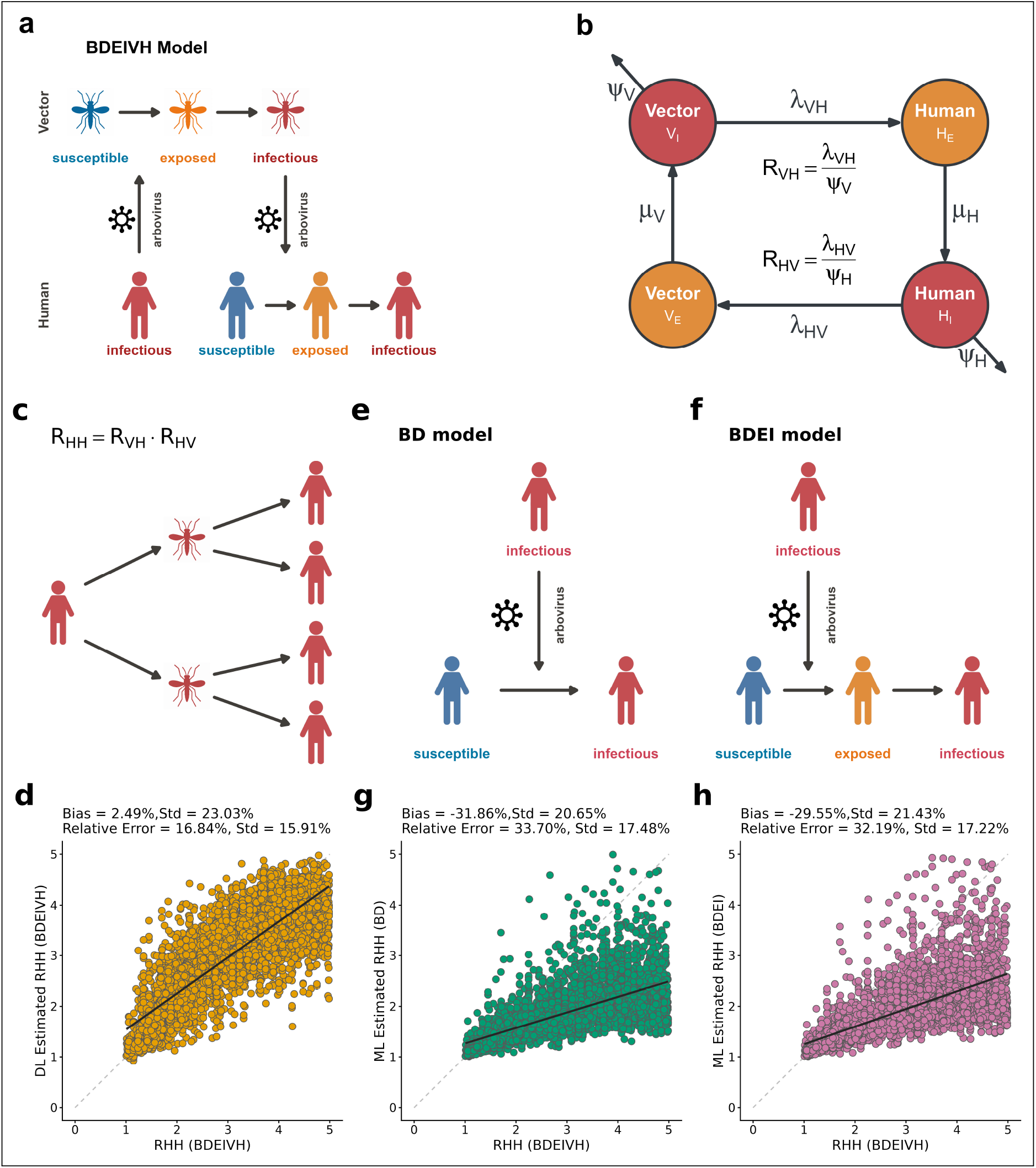
The Birth–Death Exposed–Infectious Vector–Human (BDEIVH) model. **a**. Schematic representation of arboviral transmission dynamics. b. Structure of the BDEIVH model showing the compartments, parameters and governing equations of the reproduction numbers. c. Schematic representation of the *R_HH_* parameter. d. Linear regression of the true (*R_HH_*) values (*x-axis*) of all perfectly bifurcated test trees (n = 4,000) simulated under the BDEIVH model and their predicted values (*y-axis*) using the deep-learning (DL) BDEIVH estimator. e-f. Schematic representations of the Birth–Death (BD) and Birth–Death Exposed–Infectious (BDEI) models, respectively. g-h. Linear regression of the true (*R_HH_*) values (*x-axis*) of all perfectly bifurcated test trees (n = 4,000) simulated under the BDEIVH model and their predicted values (*y-axis*) using the maximum-likelihood BD and BDEI estimators.

**Table 1:**
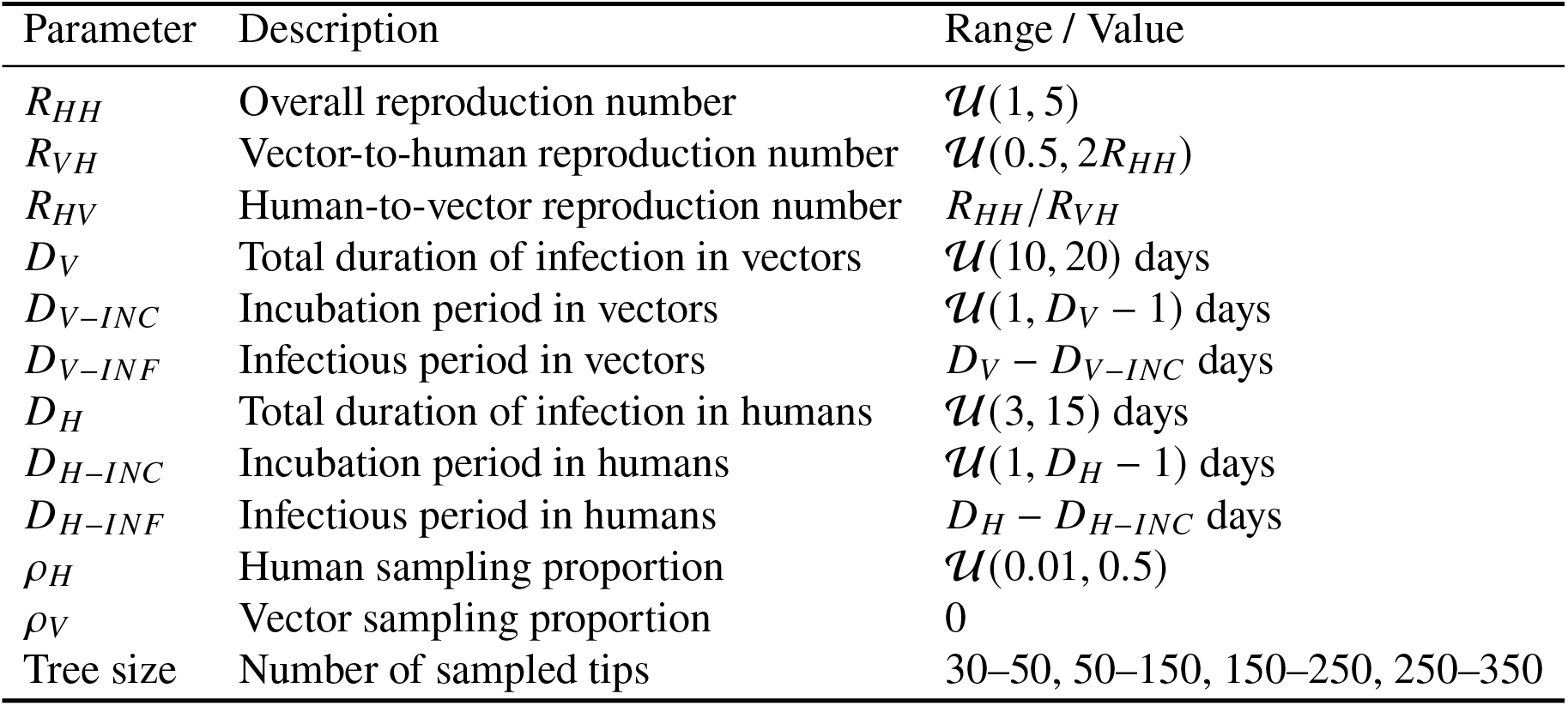
Parameter ranges used to simulate datasets under the BDEIVH model.

| Parameter | Description | Range / Value |
|---|---|---|
| *R*<sub>*HH*</sub> | Overall reproduction number | 𝒰(1, 5) |
| *R*<sub>*VH*</sub> | Vector-to-human reproduction number | 𝒰(0.5, 2*R*<sub>*HH*</sub>) |
| *R*<sub>*HV*</sub> | Human-to-vector reproduction number | *R*<sub>*HH*</sub>/*R*<sub>*VH*</sub> |
| *D*<sub>*V*</sub> | Total duration of infection in vectors | 𝒰(10, 20) days |
| *D*<sub>*V*−*INC*</sub> | Incubation period in vectors | 𝒰(1, *D*<sub>*V*</sub> − 1) days |
| *D*<sub>*V*−*INF*</sub> | Infectious period in vectors | *D*<sub>*V*</sub> − *D*<sub>*V*−*INC*</sub> days |
| *D*<sub>*H*</sub> | Total duration of infection in humans | 𝒰(3, 15) days |
| *D*<sub>*H*−*INC*</sub> | Incubation period in humans | 𝒰(1, *D*<sub>*H*</sub> − 1) days |
| *D*<sub>*H*−*INF*</sub> | Infectious period in humans | *D*<sub>*H*</sub> − *D*<sub>*H*−*INC*</sub> days |
| *ρ*<sub>*H*</sub> | Human sampling proportion | 𝒰(0.01, 0.5) |
| *ρ*<sub>*V*</sub> | Vector sampling proportion | 0 |
| Tree size | Number of sampled tips | 30–50, 50–150, 150–250, 250–350 |

### 2.2 BDEIVH Estimator Performance and Model Comparison

When applied to the 4,000 transmission trees comprising the independent test datasets across the four tree-size categories (Fig. 1d), the DL-implemented BDEIVH estimator recovered median *R_HH_* values with a small positive mean bias of 2% (SD: 23%) and a mean relative error of 17% (SD: 16%). To compare *R_HH_* inference with established likelihood-based phylodynamic approaches, the same test trees were subsequently analyzed using maximum-likelihood (ML) estimators [55, 60] of two simpler models: the birth–death (BD [52]) model (Fig. 1e) and the birth–death exposed–infectious (BDEI [53]) model (Fig. 1f). Both systematically underestimated the true *R_HH_* values used to simulate the trees (Fig. 1g–h), with mean biases of −32% (SD: 21%) for BD and −30% (SD: 21%) for BDEI. Mean relative errors were 34% (SD: 17%) and 32% (SD: 17%) for BD and BDEI, respectively, compared with 17% for the DL BDEIVH estimator. These results support that, for trees simulated under vector–human dynamics, the DL estimator reduced the systematic underestimation and relative error of *R_HH_* estimation over simpler BD models.

### 2.3 Bias in BDEIVH Predictive Performance

To more comprehensively characterize the predictive performance of the DL estimator, the true and the estimated parameters obtained for both the transmission and OROV-like datasets were compared across the four tree-size categories. Regarding relative bias, its direction and magnitude showed no overall tendency toward either over– or underestimation. mean relative biases for *R_VH_* remained relatively small, ranging from −5% to 2%, with substantial deviations from zero detected only for the two smallest categories (Table S1). A similarly heterogeneous pattern was observed for *R_HV_*, for which mean relative bias ranged from −6% to 6%, whereas *R_HH_* alternated between modest under– and overestimation across tree sizes (−5% to 4%), with significant deviations from zero in all four categories. In contrast, *D_V_* was consistently overestimated, with mean relative biases ranging from 6% to 12%, all substantially greater than zero. Estimates of *D_H_* showed a marked tree-size-dependent shift in the direction of bias, from overestimation in the two smaller categories (6% and 10%) to underestimation in the two larger categories (−10% and −9%). Likewise, *V*/*H* displayed a non-uniform pattern, although overestimation predominated, particularly in the two largest tree-size categories, where mean relative biases reached 17% and 14%. As observed for the transmission tree datasets, the OROV-like datasets (Fig. 2b) exhibited parameter– and tree-size-dependent patterns rather than pronounced unidirectional biases. The mean relative bias for *R_VH_* shifted from an overestimation of 8% in the smallest trees to modest underestimation in the larger tree-size categories (0% to −4%), whereas *R_HV_* and *R_HH_* showed comparatively small biases whose direction varied across tree sizes. Among the duration parameters, *D_V_* showed the clearest tendency toward overestimation, with positive mean relative biases across all tree-size categories (0%–12%), whereas the mean relative bias for *D_H_* ranged from −6% to 10%. Estimates of *V*/*H* were largely unbiased in the smallest trees (−5%, *P*_adj_ = 0.535) but tended toward overestimation in the three larger tree-size categories, reaching approximately 8–9% in trees over 150 tips.

**Figure 2:**
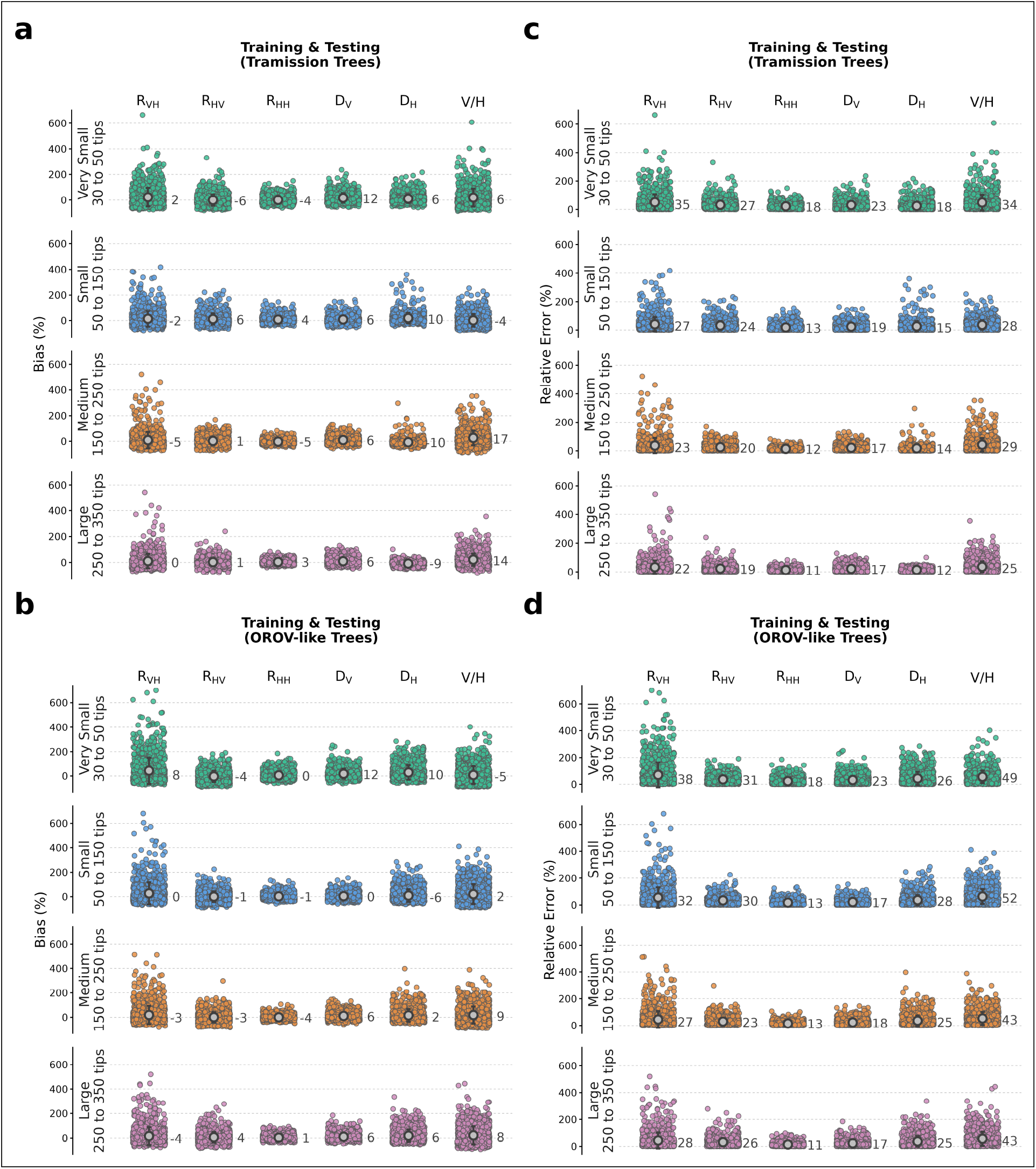
Mean Relative bias (MRB) and mean relative error (MRE) of the DL estimator of the Birth–Death Exposed–Infectious Vector–Human (BDEIVH) model. **a**, b. MRB. c, d. MRE. Results are shown for the vector-to-human (*R_VH_*) and human-to-vector (*R_HV_*) reproduction numbers, the composite human-to-human reproduction number (*R_HH_*), the duration of infection in vectors (*D_V_*) and humans (*D_H_*), and the vector-to-human ratio (*V*/*H*), across four tree-size categories and for perfectly bifurcating trees (a, c) and OROV-like trees (b, d), in which internal branches were collapsed to reproduce reduced phylogenetic resolution.

### 2.4 Relative Error in BDEIVH Predictive Performance

Regarding relative error, all six epidemiological parameters exhibited a significant reduction with increasing tree size in the transmission-tree datasets (*P* ≤ 7.77 × 10^−8^, Table S2). For *R_HH_*, for example, the median relative error decreased from 18% in trees containing 30–50 tips to 11% in the 250–350-tip category. Pairwise comparisons indicated significant improvements across successive tree-size categories up to 150–250 tips, whereas no further significant reduction was detected between the two largest categories (*P*_adj_ = 0.204, Table S3). Similar trends were observed for *R_VH_*, *R_HV_*, and *D_V_*, whereas *D_H_* showed a more continuous reduction in relative error across tree sizes and *V*/*H* exhibited a less uniform pairwise pattern. A broadly similar relationship between tree size and predictive accuracy was observed in the OROV-like datasets, although the reduction in relative error was less consistent for some parameters (Fig. 2d and Table S3 and Table S2).

### 2.5 Calibration and Precision of BDEIVH Prediction Intervals

For models trained and tested on transmission trees, empirical coverage of the 95% prediction intervals remained close to the nominal level across all parameters and tree-size categories, ranging from 93% to 97% (Fig. S2a). No systematic variation with increasing tree size was observed. A broadly similar pattern was found for OROV-like trees (Fig. S2b), for which empirical coverage also ranged from 93% to 97% for most parameters and tree-size categories. The main exception occurred in the 50–150-tip category, where lower coverage was observed for *R_VH_* (90%), *R_HV_* (88%), and *V*/*H* (91%). In contrast to the relatively stable coverage, prediction-interval width varied substantially among parameters and generally decreased with increasing tree size (Fig. S2c). For transmission trees, the relative width of the *R_HH_* prediction interval decreased from 117.3% in the 30–50-tip category to 63.0% in the 250–350-tip category. Similar reductions were observed for *R_VH_* (226.0% to 151.9%), *R_HV_* (201.4% to 140.8%), *D_H_* (124.5% to 76.6%), and *D_V_* (166.9% to 111.2%). Among the inferred parameters, *V*/*H* consistently exhibited the widest prediction intervals, although its relative width also decreased from 300.0% to 216.4% with increasing tree size. For OROV-like trees, prediction intervals were generally wider for several parameters (Fig. S2d). *R_HH_* remained comparatively well constrained, with relative width decreasing from 120.4% in the smallest trees to 66.7% in the largest. By contrast, substantially wider intervals were observed for *R_VH_*, *R_HV_*, *D_H_*, and *V*/*H*.

### 2.6 Major OROV *M*_1_*L*_2_*S*_2_ Genomic Clusters in Brazil

The time-scaled phylogeographic reconstruction of a dataset of 913 Brazilian OROV *M*_1_*L*_2_*S*_2_ complete genomes (see *Methods*) placed the root of the lineage in Amazonas in early 2014 and revealed the establishment of multiple genomic clusters in northern Brazil during 2023–2024, followed by their expansion into the Southeast and Northeast during 2024–2025, as previously described [16, 19, 25, 26]. Among the monophyletic groups identified within the phylogeny, three highly supported clusters with sufficient numbers of genomes to fall within the tree-size range supported by the DL estimator were selected: AM-I (*n* = 260) in Amazonas state (Northern region), CE (*n* = 134) in Ceará state (Northeastern region), and ES-I (*n* = 124) in Espírito Santo state (Southeastern region) (Fig. 3a). These clusters exhibited markedly different temporal patterns of circulation. AM-I was largely restricted to a relatively short period spanning from late 2023 to early 2024. ES-I persisted over a substantially longer interval extending from mid-2024 to mid-2025. Genomes belonging to the CE cluster were distributed between two temporally distinct subclusters: CE-2024 (*n* = 90), sampled from late May to late August 2024, and CE-2025 (*n* = 44), sampled from mid-February to late March 2025. AM-I, ES-I, and the two CE subclusters were further evaluated using phylogenetic summary statistics (see *Methods*), of which only CE-2025 did not meet the criteria for inclusion in the phylodynamic analysis. Thus, AM-I, CE-2024, and ES-I were retained for subsequent inference using the DL BDEIVH estimator (Table 2).

**Figure 3:**
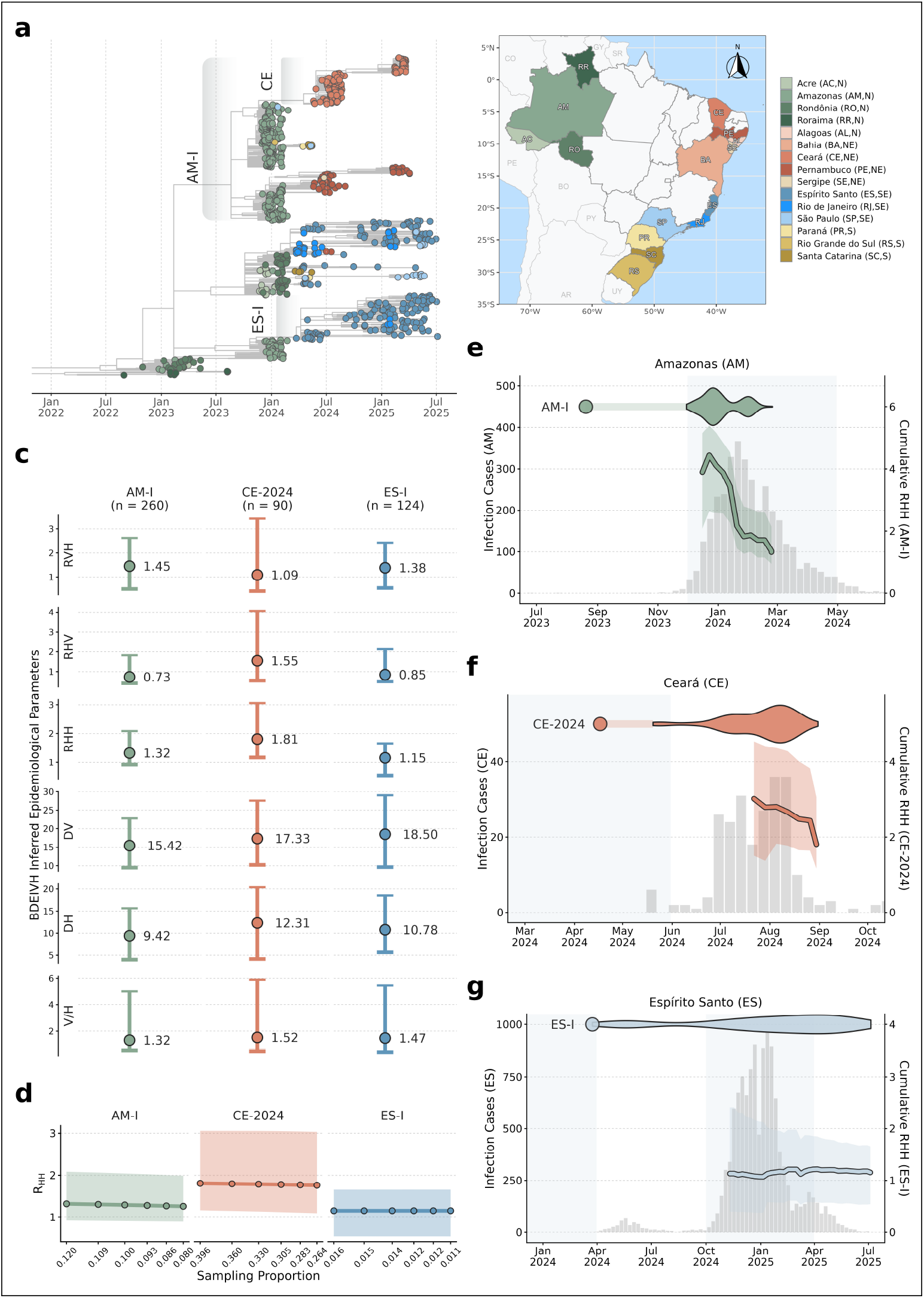
Inference of epidemiological parameters for major Oropouche virus (OROV) *M*_1_*L*_2_*S*_2_ genomic clusters in Brazil. **a**. Time-scaled phylogeny (*n* = 913) inferred from concatenated S, M, and L genome segments sampled in Brazil between 2015 and 2025 (left). Major genomic clusters are highlighted, and tips are colored according to sampling state (shown on the map on the right). b. Median estimates and 95% prediction intervals for *R_VH_*, *R_HV_*, *R_HH_*, *D_V_*, *D_H_*, and *V*/*H* across major Brazilian OROV genomic clusters. c. Sensitivity of median *R_HH_* estimates and 95% prediction intervals to sequential 10% increases in the assumed number of cases, up to 50% above baseline. d–f. Temporal evolution of cumulative *R_HH_* for AM-I (d), CE (e), and ES-I (f). Circles indicate the inferred *t*_MRCA_, transparent lines represent the interval between the inferred *t*_MRCA_ and the earliest sampled genome, and violin plots show the temporal distribution of sampled genomes. Reported cases (gray bars) and rainy seasons (light blue regions) in the corresponding states are shown in the background. *Abbreviations*: N, north; NE, Northeast; CW, Central-west; SE, Southeast; S, South.

**Table 2:** Characteristics of OROV transmission clusters selected for phylodynamic analysis.

| Cluster | aLRT | Location<br>(Probability) | Region | Sampling Range | Genomes | Sampling<br>Probability | *t*<sub>MRCA</sub><br>(Estimated) | *t*<sub>MRCA</sub><br>(Published) |
|---|---|---|---|---|---|---|---|---|
| AM-I | 0.83 | Amazonas (1.0) | North | 2023-11-30 to 2024-02-25 | 260 | 0.109 | 2023-08-20 | Jul-Oct 2023 [16] |
| CE-2024 | 0.98 | Ceará (1.0) | Northeast | 2024-05-20 to 2024-08-31 | 90 | 0.396 | 2024-04-17 | Feb-Apr 2024 [24] |
| ES-I | 0.96 | Espírito Santo (1.0) | Southeast | 2024-03-24 to 2025-07-05 | 124 | 0.016 | 2024-03-24 | Jan-Apr 2024 [19] |

*Abbreviations: aLRT, approximate likelihood-ratio test; t*<sub>MRCA</sub>*, time to the most recent common ancestor.*

### 2.7 Phylodynamic Characterization of OROV *M*_1_*L*_2_*S*_2_ Genotype in Brazil

When the DL estimators were applied to the three retained clusters, the inferred epidemiological parameters were broadly comparable (overlapping PIs) despite differences in cluster size, temporal dynamics, and outbreak setting (Fig. 3b). Median estimates of the vector-to-human reproduction number, *R_VH_*, were greater than 1 in all three clusters, ranging from 1.09 in CE-2024 to 1.45 in AM-I, with an intermediate value of 1.38 in ES-I. Greater variation was observed for the human-to-vector reproduction number, *R_HV_*, with median estimates of 0.73 in AM-I, 0.85 in ES-I, and 1.55 in CE-2024. Despite these differences in the directional components of transmission, the composite human-to-human reproduction number, *R_HH_*, had median estimates greater than 1 in all three clusters, with the highest value inferred for CE-2024 (1.81), followed by AM-I (1.32) and ES-I (1.15). The duration-related parameters showed comparatively limited variation among clusters. Median estimates of the total duration of infection in vectors, *D_V_*, ranged from 15.4 days in AM-I to 18.5 days in ES-I, with an intermediate value of 17.3 days in CE-2024. Likewise, median estimates of the total duration of infection in humans, *D_H_*, ranged from 9.42 days in AM-I to 12.3 days in CE-2024, with an estimate of 10.8 days in ES-I. The inferred vector-to-human ratio, *V*/*H*, was also similar across clusters, with median estimates of 1.32 in AM-I, 1.52 in CE-2024, and 1.47 in ES-I.

We also assessed the sensitivity of *R_HH_* inference to uncertainty in the genomic sampling proportion. Because OROV infections are likely under-ascertained, the true number of cases may exceed those reported through routine surveillance, resulting in a lower genomic sampling proportion than initially estimated. Accordingly, inference was repeated for each of the three genomic clusters after cumulatively increasing the estimated number of cases in 10% increments, up to 50% above the baseline estimate, thereby progressively reducing the assumed genomic sampling proportion (Fig. 3c). Across all three clusters, *R_HH_* estimates were only modestly affected. Median *R_HH_* decreased from 1.32 to 1.26 in AM-I (∼ 4.5%) and from 1.81 to 1.76 in CE-2024 (∼ 2.5%), whereas virtually no change was observed for ES-I (1.154 to 1.155).

### 2.8 Temporal Dynamics of *R_HH_* Across OROV Genomic Clusters

To further characterize the temporal dynamics of *R_HH_* across the three genomic clusters retained for analysis, cumulative *R_HH_* estimates were obtained by progressively slicing each phylogenetic tree and incorporating increasing amounts of information from its root toward the most recently sampled sequence (Fig. 3d–f). Starting from the inferred *t*_MRCA_, each tree was truncated at successive seven-day intervals, and *R_HH_* was re-estimated using all phylogenetic information available up to each time point. Inference was initiated once the truncated tree contained at least 30 tips, corresponding to the minimum tree size represented in the datasets used to train the DL estimators. Whereas whole-cluster analyses yielded broadly comparable *R_HH_* estimates, this cumulative approach enabled transmission dynamics to be examined in relation to the temporal accumulation of genomic information and the epidemiological context of each cluster, including the timing of genomic sampling, reported case incidence, and seasonal patterns.

AM-I (Fig. 3d and Table 2) was the largest of the three clusters (*n* = 260 genomes) and was inferred to have originated in Amazonas state in mid-2023 (*t*_MRCA_ = 20 August 2023). Genomes assigned to this cluster were sampled from late November 2023 to late February 2024 (∼ 3 months), a period that largely coincided with the rainy season in the state. AM-I accounted for more than 80% of the genomes sampled in Amazonas during this interval and comprised predominantly sequences obtained in the state capital, Manaus (∼ 70%). The concentration of genomic sampling early in the outbreak, with more than 50% of sequences collected by the end of December 2023, provided sufficient phylogenetic information to estimate cumulative *R_HH_* during the phase of case expansion in the state. Cumulative inference indicated that *R_HH_* peaked above 4.0 in late December 2023 (4.4, 95% PI: 2.6–5.4), approximately three weeks before reported case counts in Amazonas reached their peak of approximately 350 cases per epidemiological week (EW), after which *R_HH_* progressively declined until its final value of 1.3 (95% PI: 0.9–2.0).

CE-2024 (*n* = 90 genomes, Fig. 3e and Table 2) represented a subcluster within the broader CE cluster, which comprised all genomes sampled in Ceará. This subcluster was inferred to have originated in mid-May 2024 (*t*_MRCA_ = 17 April 2024), during the rainy season in the state. Nevertheless, the vast majority (> 95%) of CE-2024 genomes were sampled subsequently, between June and August 2024. Spatially, all genomes assigned to this subcluster were obtained from municipalities within the Maciço de Baturité, a humid montane enclave embedded in the semiarid region of Ceará and characterized by remnants of tropical humid forest. Reported cases in the state followed a broadly concordant temporal pattern, increasing progressively to a peak of nearly 40 cases per EW in mid-August before declining throughout September. More than 75% of CE-2024 genomes were sampled between mid-July and August 2024, such that sufficient phylogenetic information for cumulative inference became available only from late July onward, when reported case counts were already approaching their peak. At the earliest time point for which cumulative inference was possible, the median *R_HH_* was estimated at 3.0 (95% PI: 1.5–4.5) and subsequently declined as the local outbreak progressed until reaching 1.8 (95% PI: 1.2–3.0).

ES-I (Fig. 3f and Table 2) was inferred to have originated in late March 2024 (*t*_MRCA_ = 24 March 2024) and exhibited a markedly prolonged pattern of circulation, with genomes sampled across more than 40 municipalities before, during, and after the 2025 rainy season in the state. ES-I was the largest (*n* = 124 genomes) of the four genomic clusters identified in the state, substantially exceeding the sizes of the other three clusters (*n* = 6, 9, and 60) and accounting for approximately 60% of all genomes sampled in Espírito Santo. Distinct from the genomic sampling, reported cases in the state over the same period were strongly concentrated during the 2025 rainy season. Although sufficient phylogenetic information to support cumulative *R_HH_* inference became available during the rise in reported case counts, ES-I had already been detected through genomic surveillance in the state for more than six months by that time. The median cumulative *R_HH_* fluctuated only modestly, from approximately 1.0 (95% PI: 0.4–2.1) in early January to 1.2 (95% PI: 0.5–1.8) in early April, remaining relatively stable throughout the period for which inference was possible.

## 3 Discussion

Here, we introduce the Birth–Death Exposed–Infectious Vector–Human (BDEIVH) model, a novel phylodynamic framework that explicitly represents the alternating human–vector transmission cycle characteristic of arboviruses, and apply it to investigate the expansion of the OROV *M*_1_*L*_2_*S*_2_ lineage in Brazil. The model incorporates the directional reproduction numbers *R_VH_* and *R_HV_* and their product, *R_HH_*, defined as the expected number of secondary human infections generated by a single infectious human through one complete vector-mediated transmission cycle. It also describes the total durations of infection in humans (*D_H_*) and vectors (*D_V_*) and the ratio of infected vectors to infected humans (*V*/*H*). Implemented within a neural-network framework adapted from PhyloCNN [68], the BDEIVH estimator recovered key epidemiological parameters with low bias, although precision varied across parameters and tree sizes, overall improving with tree size. Performance was largely maintained when simulated trees were modified to approximate the limited phylogenetic resolution of empirical OROV data. Notably, *R_HH_* estimates were substantially more accurate than those obtained using simpler birth–death formulations applied to the same trees, illustrating how biologically informed model complexity can improve phylodynamic inference. Application to major transmission clusters in Brazil supported sustained transmission both in historically endemic regions and in areas where autochthonous OROV transmission had not been documented before the 2022–2025 outbreak.

The principal limitation of this study was the scarcity of empirical data to inform the biological assumptions underlying our simulation-based inference. In particular, limited knowledge of the extrinsic incubation period and lifespan of *C. paraensis*, presumed to be the main vector during the outbreak, prevented us from specifying separate, empirically informed ranges for the incubation and infectious periods in vectors. A further constraint was the absence of genomic sequences from infected vectors. Obtaining such data remains challenging, given reported detection frequencies of approximately one OROV-positive individual per 10,000 vectors sampled [71, 72]. Our median estimates of the infected vector-to-human ratio (*V*/*H*), ranging from 1.32 to 1.52 across the three clusters, were compatible with a substantial pool of infected vectors unrepresented in the genomic dataset. The limited biological information and exclusive reliance on human-derived sequences, when combined, constrained our ability to disentangle the incubation and infectious periods in vectors, thereby limiting the temporal detail with which we could characterize vector–human transmission. The relatively broad prediction intervals for some duration-related parameters further illustrate the uncertainty associated with these estimates.

The availability of human-derived genomic data also limited the application of our estimator. Most identified genomic clusters were relatively small, and the temporal distribution of available sequences did not necessarily reflect that of reported cases in the corresponding states. Sparse genomic sampling, together with the limited phylogenetic resolution of OROV genomic data, constrained the epidemiological information recoverable from these trees. Consequently, only a subset of the identified clusters was suitable for inference using the DL estimator. This limitation was particularly consequential for the CE-2025 subcluster, which circulated in the same geographical region as CE-2024 during a subsequent outbreak. Its inclusion would have enabled a comparison of transmission dynamics across successive outbreaks within a shared regional setting. Unlike CE-2024, CE-2025 circulated during the state’s rainy season and coincided with higher reported incidence, providing an opportunity to investigate temporal variation in transmissibility and its potential association with seasonal conditions. Such a comparison would reduce the geographical heterogeneity inherent in comparisons across regions, although the greater involvement of urban areas in the 2025 outbreak [73] would introduce ecological differences that would need to be considered. Although the 44 genomes in CE-2025 placed it within the range supported by our estimator, its highly polytomous phylogeny provided insufficient information for reliable inference, as assessed via its summary statistics.

Application of the DL estimator to major clusters of the *M*_1_*L*_2_*S*_2_ lineage in Brazil yielded broadly comparable median estimates of total infection duration in vectors and humans across the northern (AM-I), southeastern (ES-I), and northeastern (CE-2024) regions, with overlapping 95% PIs. Median estimates ranged from 15 (AM-I) to 19 days (ES-I) in vectors and from 9 (AM-I) to 12 days (CE-2024) in humans. Although empirical data on the course of OROV infection remain limited, particularly in vectors, previous studies provide plausibility for these estimates in both host types [11, 71, 74, 75]. However, our simulation-based validation revealed a tendency to overestimate duration-related parameters, warranting caution when interpreting both their absolute values and their apparent similarity across clusters.

These broadly similar estimates are compatible with transmission involving vector populations with comparable biological characteristics and human populations with similar infection kinetics. However, neither possibility can currently be evaluated directly. Entomological studies reporting OROV-positive specimens remain scarce; despite the scale of the 2022–2025 outbreak in Brazil, no published studies have documented vector detections associated with the outbreak [72]. Shared environmental features across tropical transmission settings in northern [16], northeastern [21, 24, 73], and southeastern [20, 23] Brazil are compatible with the involvement of vector populations with similar ecological characteristics, but do not establish their taxonomic identity or relative contribution to transmission. In humans, determining whether infection kinetics differed across regions would require prospective cohort studies combining well-resolved information on exposure timing with serial sampling and assays for infectious virus to estimate incubation and infectiousness periods [75]. Such comparative data are currently unavailable.

As with infection durations, estimates of the composite reproduction number, *R_HH_*, were broadly comparable across clusters, ranging from 1.15 (ES-I) to 1.81 (CE-2024), with overlapping 95% PIs. However, because *R_HH_* = *R_HV_ R_VH_*, similar composite values can arise from different combinations of the directional reproduction numbers. In AM-I and ES-I, point estimates of *R_VH_* (1.45 and 1.38, respectively) exceeded those of *R_HV_* (0.73 and 0.85, respectively), whereas CE-2024 showed the reverse pattern, with *R_HV_* = 1.55 and *R_VH_* = 1.09. These estimates suggest differences in the relative contributions of the two stages of the transmission cycle. Assuming comparable vector competence, per-vector biting rates, and human infectiousness across settings, the higher *R_HV_* estimated for CE-2024 could reflect greater exposure of infectious humans to susceptible vectors. Such exposure could also contribute to a higher ratio of infected vectors to infected humans (*V*/*H*), a quantity distinct from the overall vector-to-human abundance ratio. Consistent with this hypothesis, the median estimated *V*/*H* was slightly higher in CE-2024 (1.52) than in AM-I (1.32) and ES-I (1.47). Nevertheless, *V*/*H* was the least reliably estimated of the six quantities in our validation analyses, and these modest differences therefore provide limited support for this hypothesis.

An additional finding from our analyses was the robustness of *R_HH_* estimation to variation in the sampling proportion, with negligible changes in estimates when the assumed number of cases associated with transmission was increased by up to 50%. This result is particularly relevant given evidence that epidemiological surveillance may fail to detect a substantial fraction of OROV infections [73, 76–79]. Underascertainment may arise from the nonspecific clinical presentation of OROF, which overlaps with that of dengue and other febrile illnesses [4] in settings where multiple arboviruses circulate [21, 80], as well as from asymptomatic or oligosymptomatic infections [73]. A population-based serosurvey conducted after the outbreak in Baturité, Ceará, found that approximately 75% of seropositive participants reported no history of Oropouche fever, consistent with a substantial burden of asymptomatic, oligosymptomatic, or otherwise unrecognized infections [73].

To complement the constant *R_HH_* estimates obtained for each phylogeny and better characterize the epidemiological dynamics of individual transmission clusters, we applied a systematic temporal sub sampling approach, advancing from the root of each tree in seven-day increments to assess variation in *R_HH_* over time. For AM-I, the temporal distribution of sampled genomes closely mirrored that of reported cases, enabling estimation of *R_HH_* during rapid epidemic growth. Estimates ranged from 4.10 (95% PI: 2.6–5.1) to 4.43 (95% PI: 2.6–5.4) in epidemiological week (EW) 51 of 2023 (17–23 December). The estimated peak in *R_HH_* preceded the peak in reported cases by approximately three weeks. This lag may reflect the continued accumulation of infections following peak transmission potential, compounded by delays associated with both incubation periods and case reporting. The subsequent peak in statewide case counts may also reflect the expansion of other transmission clusters co-circulating in Amazonas [16]. Using weekly reported case counts for Brazil, Rodriguez-Morales *et al.* [81] estimated *R*_0_ at 2.87 (95% CI: 2.36–3.55) using an exponential-growth method and 3.79 (95% CI: 3.30–4.35) using a maximum-likelihood method during the early growth phase of the first epidemic wave (EW 45–50 of 2023), when approximately 70% of reported cases were concentrated in Amazonas. The substantial overlap between confidence intervals of those estimates suggests broad compatibility between the epidemiological and phylodynamic findings. Nevertheless, differences in geographical scope, temporal aggregation, number of genetic clusters considered, and parameter definition warrant caution in direct comparisons between studies.

For CE-2024, the highest *R_HH_* estimate obtained through our cumulative approach was 3.0 (95% PI: 1.5–4.5), corresponding to epidemiological week (EW) 29 of 2024 (16–22 July). This median estimate was lower than that obtained for AM-I, however, overlapped substantially. Moreover, direct comparisons warrant caution because the two clusters circulated in distinct geographical and seasonal contexts. While the spread of the AM-I clade between late 2023 and early 2024 coincided with the rainy season in Amazonas, most reported cases in Ceará in 2024 occurred during July and August, outside the state’s rainy season. Regardless of its potential relative to the endemic region, these findings suggest substantial transmission potential for OROV in a non-endemic northeastern region, even outside the optimal period of rainy season.

For ES-I, the temporal distribution of genomic sampling was poorly aligned with that of reported cases, with sequences collected across periods of markedly different epidemiological activity rather than being concentrated around the epidemic peak. Consequently, although sufficient phylogenetic information for cumulative *R_HH_* inference became available during the rapid increase in reported cases, these estimates incorporated genomic information accumulated over a prolonged and epidemiologically heterogeneous period, including months of substantially lower reported incidence. This temporal heterogeneity may have obscured changes in transmission intensity during the epidemic expansion, potentially contributing to the relatively stable *R_HH_* estimates observed for ES-I. Interpretation is further complicated by the co-circulation of multiple transmission clusters in Espírito Santo [19], whose relative contributions to statewide incidence and temporal patterns of expansion remain unresolved. Using Bayesian inference based on daily case counts, Delatorre *et al.* [20] estimated an instantaneous reproduction number (*R_t_*) of approximately 2.5 between epidemiological weeks (EWs) 18 and 27 of 2024, reaching a peak of approximately 3.0 in EW 26. However, this earlier period of elevated transmission was not captured by our cumulative analysis, precluding a direct comparison between the epidemiological and phylodynamic estimates during that phase of the outbreak.

Future development of the BDEIVH framework should include extending inference to larger phylogenies and broadening the simulation datasets used for training. Training on trees with more than 350 tips would enable the analysis of larger transmission clusters, including those encountered in other arboviral systems, while allowing us to assess whether the observed gains in predictive accuracy persist beyond the current size range. Increasing the number of training trees may also improve performance by providing denser coverage of parameter space. For several parameters, however, improvements were modest across the larger tree-size categories. This pattern could reflect limitations in the information conveyed by the phylogenies, the training data, or the estimator. Evaluating training dataset size and phylogeny size jointly would help distinguish these possibilities and establish whether additional simulations improve inference where larger trees offer limited gains.

A further extension would be to introduce a skyline formulation [53], allowing key parameters to vary across successive time intervals within a transmission cluster. This would enable the framework to characterize temporal variation in transmissibility that may be obscured by estimates averaged over the cluster’s history. Such flexibility would be particularly relevant to arboviral outbreaks, where transmission dynamics can change with seasonality [82–85], vector abundance [85–87], population susceptibility [88, 89], and control measures [90–92]. The biological basis of the model could also be strengthened as more precise estimates of vector– and host-specific parameters become available. Where OROV-specific evidence remains scarce, experimental and epidemiological studies of related arboviruses could help inform plausible ranges for the extrinsic incubation period and the infectious periods in vectors and vertebrate hosts. The applicability of these estimates would nevertheless require careful assessment, given potential differences among virus–vector–host systems and the environmental conditions under which they were obtained.

Finally, a further extension would be to integrate reported case counts with phylogenetic data within the BDEIVH framework. Particle filtering offers a way to combine these complementary sources of information by representing plausible epidemic trajectories as a collection of particles, each corresponding to a distinct realization of the underlying epidemic process in the likelihood-based context [93–95]. In the DL context, case counts could be represented as additional features associated with input tree data. For OROV, this approach could help resolve temporal variation in *R_HH_* and better constrain infection dynamics in human and vector populations. The observation and simulation models would need to accommodate under reporting, reporting delays, and changes in surveillance practices, while ensuring that case counts correspond to the population represented by the phylogeny. Developing this extension would also require an explicit representation of the relationship between case reporting and genomic sampling.

This study advances arboviral phylodynamics by combining for the first time an explicit representation of the alternating vertebrate–vector–vertebrate transmission cycle with a DL approach that makes parameter inference computationally tractable. Applied to OROV, the estimator yielded epidemiologically plausible estimates of transmission parameters across distinct Brazilian regions. Its relevance extends beyond OROV, providing a methodological foundation for studying other arboviruses, subject to appropriate biological parameterization and validation. Future work should prioritize expanding the size and diversity of training datasets, incorporating time-varying transmission parameters, refining biological parameterization, and integrating epidemiological data into the inference framework. These developments would strengthen our ability to investigate how ecological conditions shape arboviral transmission and the resulting burden of disease.

## 4 Materials and Methods

### 4.1 The Birth–Death Exposed–Infectious Vector–Humans (BDEIVH) model

To capture the dynamics of arboviral transmission in urban cycles (Fig. 1a) and enable their subsequent integration into a DL framework, we developed the Birth–Death Exposed–Infectious Vector–Human (BDEIVH) model (Fig. 1b), a MTBD process that explicitly represents pathogen transmission between vector and human hosts. The model comprises four epidemiological states: exposed vectors (*V_E_*), infectious vectors (*V_I_*), exposed humans (*H_E_*), and infectious humans (*H_I_*). Exposed individuals are infected but not yet capable of onward transmission, whereas infectious individuals transmit infection between hosts at rates *λ_VH_* (vector-to-human) and *λ_HV_* (human-to-vector). Progression from the exposed to the infectious state occurs at rates *μ_V_* and *μ_H_*, corresponding to the inverse of the mean extrinsic (vector) and intrinsic (human) incubation periods. Infectious vectors and humans are removed from the transmission process at rates *ψ_V_* and *ψ_H_*, representing the inverse of their mean infectious periods (for definitions and interpretations of phylodynamic parameters see [96, 97]). Human infections are sampled upon removal with probability *ρ_H_*, whereas vector infections are sampled with probability *ρ_V_*.

The model is parameterized by two directional reproduction numbers:

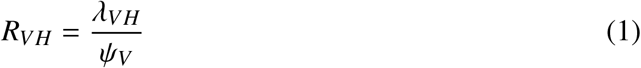

and

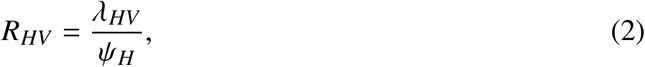

where *R_VH_* denotes the expected number of human infections generated by a single infectious vector, and *R_HV_* denotes the expected number of vector infections generated by a single infectious human. We also define *R_HH_* (Fig. 1c), a composite human-to-human reproduction number associated with a complete transmission cycle involving both humans and vectors, which is defined as:

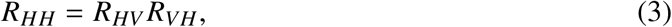

which quantifies the expected number of secondary human infections generated by a single infectious human through one complete vector-mediated transmission cycle.

### 4.2 Simulation Parameters

The DL-implemented BDEIVH estimator was trained using tree datasets simulated under parameters informed by OROV biology in both vector and human hosts, as well as by genomic surveillance data from Brazil (Table 1). Pinheiro *et al*. [71] reported an extrinsic incubation period of 6–12 days. As for the infectious period in vectors, Roberts *et al*. [74] observed approximately 50% mortality among field-collected vectors after 10–11 days under laboratory conditions, with mortality reaching 100% ≈ 6 days later. The age of the individuals at the time of capture was unknown, limiting the extent to which these observations can be interpreted as estimates of total adult longevity. Given the limited empirical information available to independently parameterize the incubation (*D_V_*_−_*_INC_*) and infectious (*D_V_*_−_*_INF_*) periods, these components were jointly represented as the total duration of infection in vectors (*D_V_*), ranging uniformly between 10 and 20 days, from which the incubation and infectious periods were randomly sampled.

The intrinsic incubation period was initially reported by Pinheiro *et al*. [11] to range from 4–8 days, whereas the infectious period generally lasted 2–5 days, although durations of up to 10 days were also observed. The median incubation period for OROV was estimated at 3.2 days, with the 5th and 95th percentiles of the incubation-period distribution estimated at approximately 1.1 and 9.7 days, respectively [75]. Consistent with the parameterization adopted for vectors, the incubation (*D_H_*_−_*_INC_*) and infectious (*D_H_*_−_*_INF_*) periods in humans were jointly represented as the total duration of infection, *D_H_*, with a uniform range of 3 to 15 days.

The vector-to-human and human-to-vector reproduction numbers, *R_VH_* and *R_HV_*, respectively, were parameterized such that their product corresponded to *R_HH_*, which was sampled uniformly between 1 and 5. This range was informed by two OROV studies in Brazil. First, a nationwide analysis of the 2023–2024 epidemic peaks estimated median *R_t_* values ranging from 2.87 to 4.26, with the upper bounds of the corresponding 95% confidence intervals reaching 4.73 [81]. Second, an analysis conducted in the state of Espírito Santo inferred a median *R_t_* of approximately 2.5, with peak values reaching 3.0 [20]. For each simulation, *R_HH_* was first sampled from its predefined range, after which *R_VH_* was sampled uniformly between 0.5 and 2*R_HH_*. The corresponding value of *R_HV_* was then constrained by the relationship *R_HV_* = *R_HH_*/*R_VH_*. Therefore, both *R_VH_* and *R_HV_* were allowed to vary within an overall range of 0.5 to 10, while still satisfying the relation *R_VH_ R_HV_* = *R_HH_*.

The last two parameters were informed by genomic clusters previously described in Brazil [16, 20, 21]. The sampling proportion in humans, *ρ_H_*, was allowed to vary between 1% and 50%, while the vector sampling proportion, *ρ_V_*, was fixed at zero since no OROV genome sequences derived from vector samples were available. Regarding tree size, four categories were considered: (*i*) 30–50, (*ii*) 50–150, (*iii*) 150–250, and (*iv*) 250–350 tips, enabling the evaluation of the effect of tree size on the predictive performance of the DL estimator.

### 4.3 Tree Simulation

Tree datasets were simulated using treesimulator v.0.2.3 [60], available at https://github.com/ evolbioinfo/treesimulator. For each of the four tree-size categories, three independent datasets were generated: a training set comprising *n* = 100,000 trees, a calibration set comprising *n* = 2,000 trees, and a test set comprising *n* = 1,000 trees (see *Neural Network Architecture* and *Estimator Inference Uncertainty*) (Fig. S1a–j). In addition to the parameters described above, the ratio between the numbers of vector and human hosts represented in each tree (*V*/*H*) was recorded. Thus, twelve quantities were retained for each simulated tree: *R_VH_*, *R_HV_*, *R_HH_*, *D_V_*, *D_V_*_−_*_INC_*, *D_V_*_−_*_INF_*, *D_H_*, *D_H_*_−_*_INC_*, *D_H_*_−_*_INF_*, *V*/*H*, *ρ_H_*, and the number of tips.

### 4.4 Generation of OROV-like Datasets

To generate phylogenies that more closely resemble those observed empirically, we applied a transformation procedure analogous to that described by Xie *et al*. [70] to training, test, and calibration tree sets, with modifications tailored to OROV. Briefly, idealized time-scaled fully bifurcated trees were transformed into OROV-like phylogenies by assigning a number of nucleotide substitutions to each branch according to a Poisson process parameterized by (*i*) the branch duration in years, (*ii*) the estimated OROV evolutionary rate (1.2 × 10^−3^, *subs*/*site*/*year* SD = 7.5 × 10^−5^) [16], and (*iii*) the complete genome length of the three segments (L, M, and S) concatenated (11, 874 nucleotides) [3]. Branches assigned less than one substitution were subsequently collapsed, reproducing the loss of phylogenetic resolution that occurs when closely related viral genomes are genetically indistinguishable. In contrast to the original procedure [70], the resulting polytomies were retained rather than arbitrarily resolved, thereby avoiding the introduction of branching relationships unsupported by the simulated genetic data. The transformed trees were subsequently re-dated using LSD2 v2.4.4 [98] to generate time-scaled OROV-like phylogenies that were then used to train the DL estimator.

### 4.5 Phylogenetic Tree Encoding

The simulated trees were encoded following the principles introduced by PhyloCNN [68], using a windowing approach in which the local neighborhood of each internal node and leaf is represented by a set of tree-derived features. Given the comparable performance obtained when encoding two or three hierarchical levels of node neighborhoods, we restricted the representation to two levels (No-context and 1-neighborhood), thereby reducing the dimensionality of the encoded data [68]. Accordingly, 10 features were retained from the original PhyloCNN encoding. From the No-context level: (*i*) sampling probability of the tree (informed alongside the supplied empirical trees); (*ii*) time from the focal node to the root; (*iii*) time from the focal node to the present; (*iv*) time from the focal node to its parent; and (*v*) number of descendant leaves. From the 1-neighborhood level: (*vi*) time from the focal node to its grandparent; (*vii*) time to the child with the smaller number of descendant leaves; (viii) time to the child with the larger number of descendant leaves; (*ix*) number of leaves descending from the child with the smaller number of descendant leaves; (*x*) number of leaves descending from the child with the larger number of descendant leaves. When polytomies were present, either in the OROV-like trees or in the empirical tree, they were retained. To explicitly incorporate information on polytomies into the tree representation, two additional features were recorded for each focal node: (*xi*) the median number of descendant leaves subtended by each of its direct children; and (*xii*) the standard deviation of the number of descendant leaves subtended by each of its direct children.

### 4.6 Neural Network Architecture

The DL estimator was adapted from PhyloCNN [68] and implemented in Python using TensorFlow v.2.13.0 [99] and Keras v.2.13.1 [100]. Each encoded phylogeny was represented as two separate input channels, corresponding to leaves and internal nodes. Within each channel, leaves and internal nodes were independently ordered according to their distance (in terms of number of generations) from the root and zero-padded to a maximum of 350 rows, resulting in an input tensor of dimensions 350 × 12 × 2, where the 12 features correspond to the tree-derived attributes (see *Tree Encoding*) and the two channels distinguish leaves from internal nodes. The convolutional component followed a depthwise separable architecture. First, a depthwise convolutional layer with 32 filters and a 1 × 12 kernel was applied using two groups, such that the leaf and internal-node channels were processed separately. This was followed by a pointwise convolutional layer comprising 32 1 × 1 filters, which combined information across the two channels, and by an additional nonlinear convolutional layer with 32 1 × 1 filters. Each convolutional layer used an exponential linear unit (ELU) activation function and was followed by batch normalization. The resulting 32 feature maps were subsequently summarized by global average pooling and passed to a feed-forward neural network comprising four fully connected layers with 64, 32, 16, and 8 neurons, respectively, each using ELU activation. Finally, the output head generated 18 (3 × 6) intermediate values, corresponding to three outputs for each of the six epidemiological parameters targeted for inference (*R_VH_*, *R_HV_*, *R_HH_*, *D_V_*, *D_H_*, and *V*/*H*). These outputs were transformed into the 2.5%, 50%, and 97.5% conditional quantiles (see *Estimator Inference Uncertainty*).

### 4.7 Estimator Inference Uncertainty

The network was trained using the *pinball loss* [101], allowing the estimation of the 2.5%, 50%, and 97.5% conditional quantiles for each epidemiological parameter. The pinball loss function for a quantile *τ* was defined as:

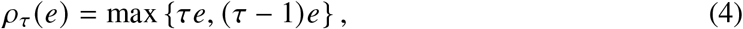

where 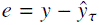 and *τ* ∈ {0.025, 0.5, 0.975}. The predicted quantiles were made positive and structurally dependent rather than treated as unconstrained independent outputs. Given three raw network outputs, *z*_1_, *z*_2_, and *z*_3_, the lower, median, and upper quantiles were defined as:

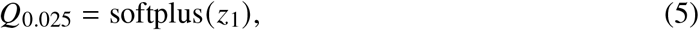

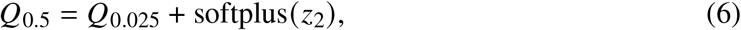

and

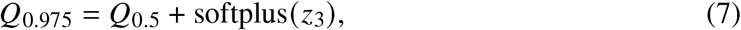

thereby guaranteeing 0 ≤ *Q*_0.025_ ≤ *Q*_0.5_ ≤ *Q*_0.975_ and preventing quantile crossing. Finally, the resulting prediction intervals were calibrated using *conformalized quantile regression* (CQR) [102] on an independent calibration dataset. For each calibration observation, the conformity score was calculated as:

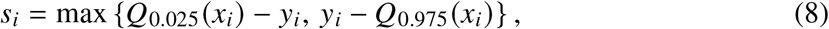

and the corresponding conformal correction, *q*^, was subsequently used to obtain the calibrated prediction interval:

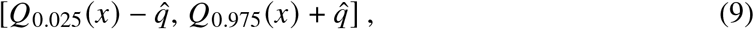

thereby improving the empirical coverage of the estimated uncertainty intervals.

### 4.8 Estimator Predictive Performance

To evaluate the performance of the DL estimator, we compared its parameter predictions with the corresponding true values in independent test datasets (n = 1,000 per tree-size category) comprising trees sampled from the same size ranges and represented using the same encoding procedures as those used for training. Estimator performance was quantified using the mean relative bias (MRB) and mean relative error (MRE), as proposed by Voznica *et al.* [56]:

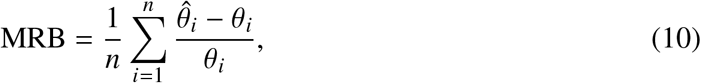

and

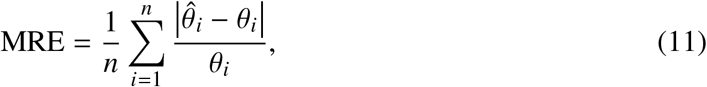

where *n* denotes the number of trees in the test dataset, *θ_i_* is the true value of a given epidemiological parameter used to simulate the *i*th tree, and 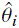 is the corresponding estimate produced by the estimator.

### 4.9 Genomic Dataset and Genotyping

To identify major genomic clusters within the OROV *M*_1_*L*_2_*S*_2_ lineage in Brazil, all publicly available genomes (*n* = 980) meeting the following criteria were downloaded from the GISAID EpiArbo platform (https://gisaid.org/): (*i*) complete (L, M, and S segments); (*ii*) high-quality (≤ 5% unidentified positions); (*iii*) collected in the country between 2015, when the earliest sequence assigned to this lineage was sampled [103], and 2025; (*iv*) accompanied by complete sampling dates (day, month, and year); and (*v*) sampling location specified at state level. All segments were genotyped following the methodology and nomenclature proposed by Naveca *et al.* [16]. Datasets were aligned using MAFFT v7.526 [104], and the resulting alignments were manually curated using Aliview v1.32 [105] when necessary. Maximum-likelihood (ML) phylogenetic trees were inferred independently for the L, M, and S segments using IQ-TREE v3.1.3 [106]. The best-fitting nucleotide substitution model for each segment was selected using ModelFinder [107], yielding TIM3+F+I+G4 for the L and M segments and K3Pu+F+I+G4 for the S segment. Branch support was assessed using the Shimodaira–Hasegawa-like approximate likelihood-ratio test (SH-aLRT) with 1,000 replicates [108]. Genotypes were defined based on phylogenetic clustering with significant branch support (SH-aLRT values ≥ 0.9).

### 4.10 Spatiotemporal Dynamics

All genomes sampled in Brazil during the study period (*n* = 980) were classified as belonging to the *M*_1_*L*_2_*S*_2_ lineage. Their genome segments were concatenated into a single alignment comprising 11,874 nucleotide positions, from which a maximum-likelihood (ML) phylogenetic tree was inferred using the TIM3+F+I+G4 nucleotide substitution model selected by ModelFinder [107], following the previously described inference procedure. The temporal signal of the resulting ML phylogeny was assessed through a root-to-tip regression analysis using TempEst v1.5.3 [109]. Sequences with residuals outside *Q*_1_ − 1.5 × IQR and *Q*_3_ + 1.5 × IQR were considered outliers and excluded, as they are likely the result of incorrect date assignment or sequencing errors. Following removal of temporal outliers, a new ML phylogeny was constructed with the final dataset (*n* = 913) and time-scaled using LSD2 v2.4.4 [98]. Ancestral geographic states were reconstructed across the tree nodes using PastML v.1.9.51 [110] under the F81 model of character evolution and the marginal posterior probabilities approximation (MPPA) method. Sequences were assigned discrete geographic states according to their state of sampling in Brazil.

### 4.11 Genomic Cluster Selection

To be retained for phylodynamic analyses, genomic clusters were required to meet the following criteria: (*i*) strong statistical support (SH-aLRT ≥ 0.80); (*ii*) between 30 and 350 genomes, matching the range of tree sizes used to train the DL estimator; (*iii*) reconstruction of the ancestral node with high confidence (marginal probability ≥ 0.90); and (*iv*) concordance between the inferred ancestral state and the sampling location of sequences within the cluster, with ≥ 95% of sequences sampled from the same state, after excluding those from other locations that formed supported clusters (SH-aLRT ≥ 0.90). Additionally, to ensure congruence between the OROV-like training tree set and the selected empirical clusters, both were evaluated using phylogenetic summary statistics following Saulnier *et al.* [111]. The clusters for which more than one statistic deviated by more than three standard deviations from the corresponding training set were excluded from subsequent analyses.

For each retained genomic cluster, the number of associated cases was estimated by scaling the total number of reported cases according to the proportion of genomes assigned to the cluster:

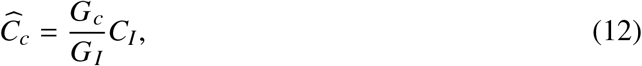

where *G_c_* is the number of genomes in cluster *c*, and *G_I_* and *C_I_* are, respectively, the total numbers of genomes and reported cases from the corresponding location during the interval between the cluster *t*_MRCA_ and its most recently sampled sequence. The genomic sampling probability was then calculated as:

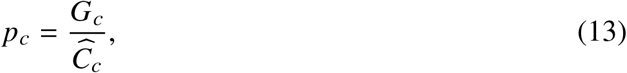

where *p_c_* represents the proportion of estimated cases represented by sampled genomes.

### 4.12 Statistical Analysis

To evaluate whether inference accuracy improved with increasing tree size, relative errors were compared across the four ordered tree-size categories using the one-sided Jonckheere–Terpstra test [112, 113], with the alternative hypothesis specifying a decreasing trend in relative error as tree size increased. Pairwise differences in relative error among tree-size categories were further assessed using Dunn’s test [114], with *P*-values adjusted for multiple comparisons using the Holm method [115]. Potential systematic bias in parameter estimation was evaluated independently within each tree-size category using two-sided Wilcoxon signed-rank tests [116], with the null hypothesis that the mean relative bias was equal to zero. For each parameter, *P*-values obtained across the four tree-size categories were adjusted for multiple testing using the Holm method [115].

### 4.13 Epidemiological and Spatial Data

Epidemiological data on the number of OROV infections in Brazil between 2023 and 2025 were obtained from the Ministry of Health (https://www.gov.br/saude/pt-br/assuntos/ saude-de-a-a-z/o/oropouche/painel-epidemiologico). The boundaries of Brazilian states were obtained from the Brazilian Institute of Geography and Statistics (https://www.ibge.gov.br/en/), using the geobr R package [117].

### 4.14 Data Availability

The Oropouche virus (OROV) genome sequences and associated metadata used in this study were retrieved from the GISAID EpiArbo database (https://gisaid.org/), being accessible through the EPI_SET_261117qg identifier (https://doi.org/10.55876/gis8.261117qg). Statistical visualizations were generated in R using ggplot2 v.4.0.3 [118], whereas phylogenetic trees were visualized using ggtree v.4.2.0 [119]. Phylogenetic tree parsing and manipulation were performed using ETE3 v.3.1.3 [120]. The Snakemake [121] pipelines, source code for training, test and calibration dataset generation, and tree encoding, as well as Jupyter notebooks used for estimator training and testing, and source code for epidemiological parameter inference with DL– and maximum-likelihood-based estimators are available on GitHub at https://github.com/modpath/bdeivh. The repository also contains the newick files with genomic clusters analyzed in this study, as well as the pretrained BDEIVH DL models. The BDEIVH estimators are also available as a command-line program and a Python 3 package via PyPi (bdeivh), and via Docker/Singularity (evolbioinfo/bdeivh).

## Acknowledgments

We gratefully acknowledge all data contributors, *i.e.*, the authors and originating laboratories responsible for obtaining the specimens, as well as the submitting laboratories responsible for generating the genetic sequences and associated metadata and for sharing them through the GISAID Initiative. We thank the Ceará State Health Department, particularly its epidemiological surveillance, vector control, and environmental health teams, and especially the Ceará Central Public Health Laboratory (LACEN-CE). We also thank the Baturité Municipal Government and Municipal Health Department, as well as the residents of the Maciço de Baturité region for their outstanding collaboration. We equally acknowledge *Institut Pasteur* for providing the computational infrastructure and server resources used in this study. I.A. was funded by the *Fundação Carlos Chagas de Amparo à Pesquisa do Estado do Rio de Janeiro* – FAPERJ (grant SEI-260003/019669/2022), and the Make out Planet Great Again (MOPGA) program (ID N^o^ 186804S). G.B. is supported by the *Conselho Nacional de Desenvolvimento Científico e Tecnológico* – CNPq through the productivity research fellowship (grant N° 304048/2024-0) and by FAPERJ through the *Cientista do Nosso Estado* (CNE) fellowship (grant N° E-26/200.112/2026). This work received funding from the Inception program (*Investissement d’Avenir* grant ANR-16-CONV-0005 awarded to S.D.), from a project grant from the *Agence Nationale de Recherche* AAPG2024 (project TrAM awarded to S.D.), and from a project grant DEELOGENY from the *Agence Nationale de Recherche* (project ANR-23-CE45-0027 awarded to A.Z.). F.G.N. is supported by the Fundação de Amparo à Pesquisa do Estado do Amazonas (FAPEAM) through Call 04/2022/FIOCRUZ/FAPEAM/FAPERO – *Inovação na Amazônia*, Call 023/2022 – *Iniciativa Amazônia +10*, the *Rede Genômica de Vigilância em Saúde do Estado do Amazonas* (REGESAM; Resolutions 002/2008, 007/2018, and 005/2019 – PRÓ-ESTADO/FAPEAM), and by the Conselho Nacional de Desenvolvimento Científico e Tecnológico (CNPq) through the *Institutos Nacionais de Ciência e Tecnologia* (INCT-VER) and Call CNPq/MCTI 10/2023 – Universal 2023, Track B – Consolidated Research Groups (grant 421620/2023-4). E.D. is supported by the Fundação de Amparo à Pesquisa e Inovação do Espírito Santo (FAPES).

## Author Contributions

I.A., A.Z., and S.D. conceived and designed the study, performed the bioinformatic analyses and drafted the original manuscript. I.A., A.Z., S.D., and G.B. contributed to data analysis. A.Z., S.D., G.B., F.G.N., A.S.L.M and E.D. contributed to fund acquisition. F.G.N., A.S.L.M and E.D. contributed to sample collection and the generation of genomic data. All authors critically reviewed and approved the final version of the manuscript.

## Supplementary Figures

**Figure S1:**
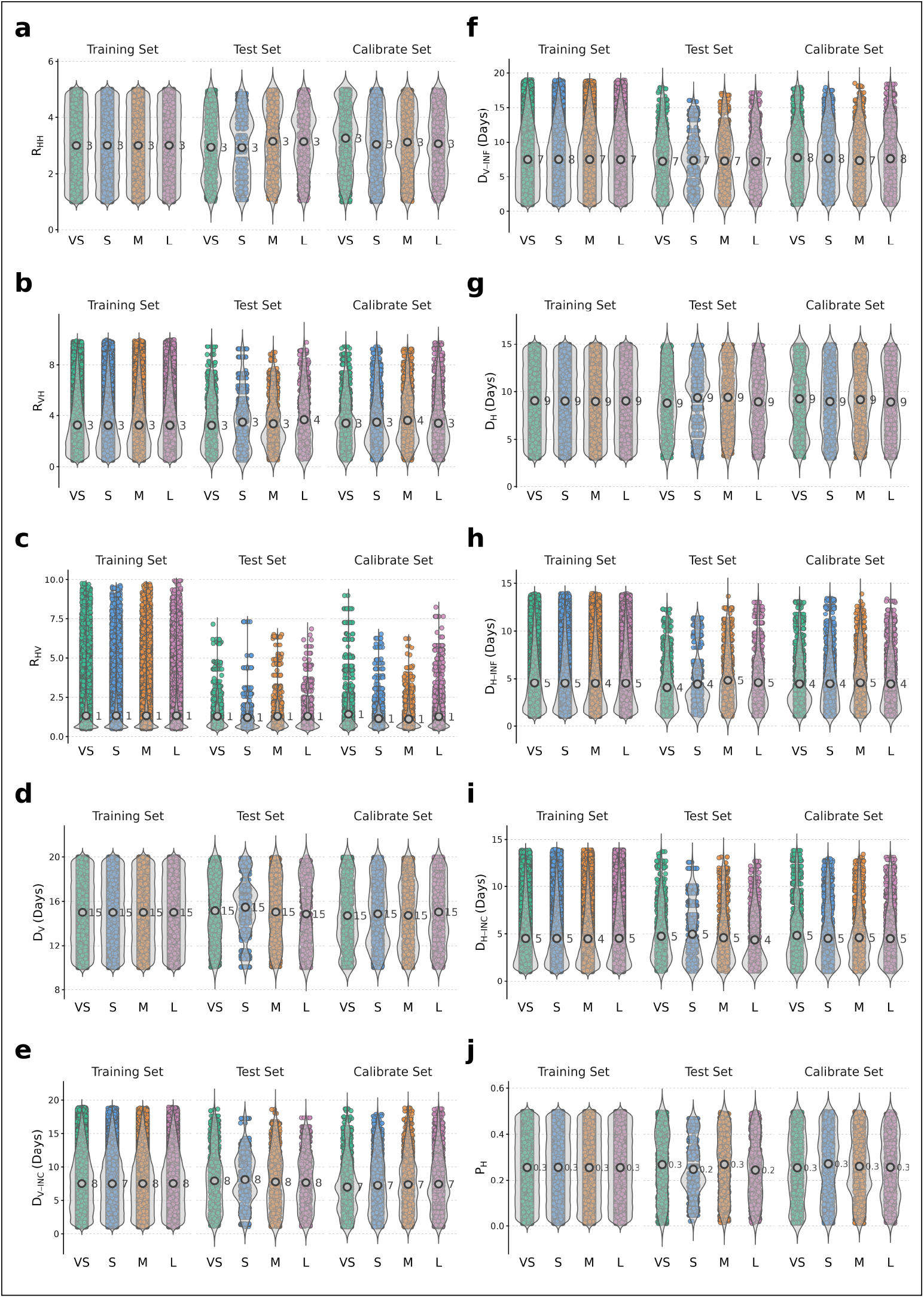
Distribution of sampled parameter values across simulated tree datasets. The figure shows the distributions of parameter values sampled to generate the training (*n* = 100,000), test (*n* = 1,000), and calibration (*n* = 2,000) datasets for *R_HH_* (a), *R_VH_* (b), *R_HV_* (c), *D_V_* (d), *D_V_*_-_*_I NC_* (e), *D_V_*_-_*_I NF_* (f), *D_H_* (g), *D_H_*_-_*_I NC_* (h), *D_H_*_-_*_I NF_* (i), and *ρ_H_* (j). Distributions are shown separately for four tree-size categories: VS (very small; 30–50 tips), S (small; 50–150 tips), M (medium; 150–250 tips), and L (large; 250–350 tips), with colors indicating the corresponding dataset. Each point represents an individual sampled parameter value (x-axis), while the superimposed violin plot summarizes the distribution of sampled values within each dataset and tree-size category. The median value for each distribution of values is also annotated in the plot.

**Figure S2:**
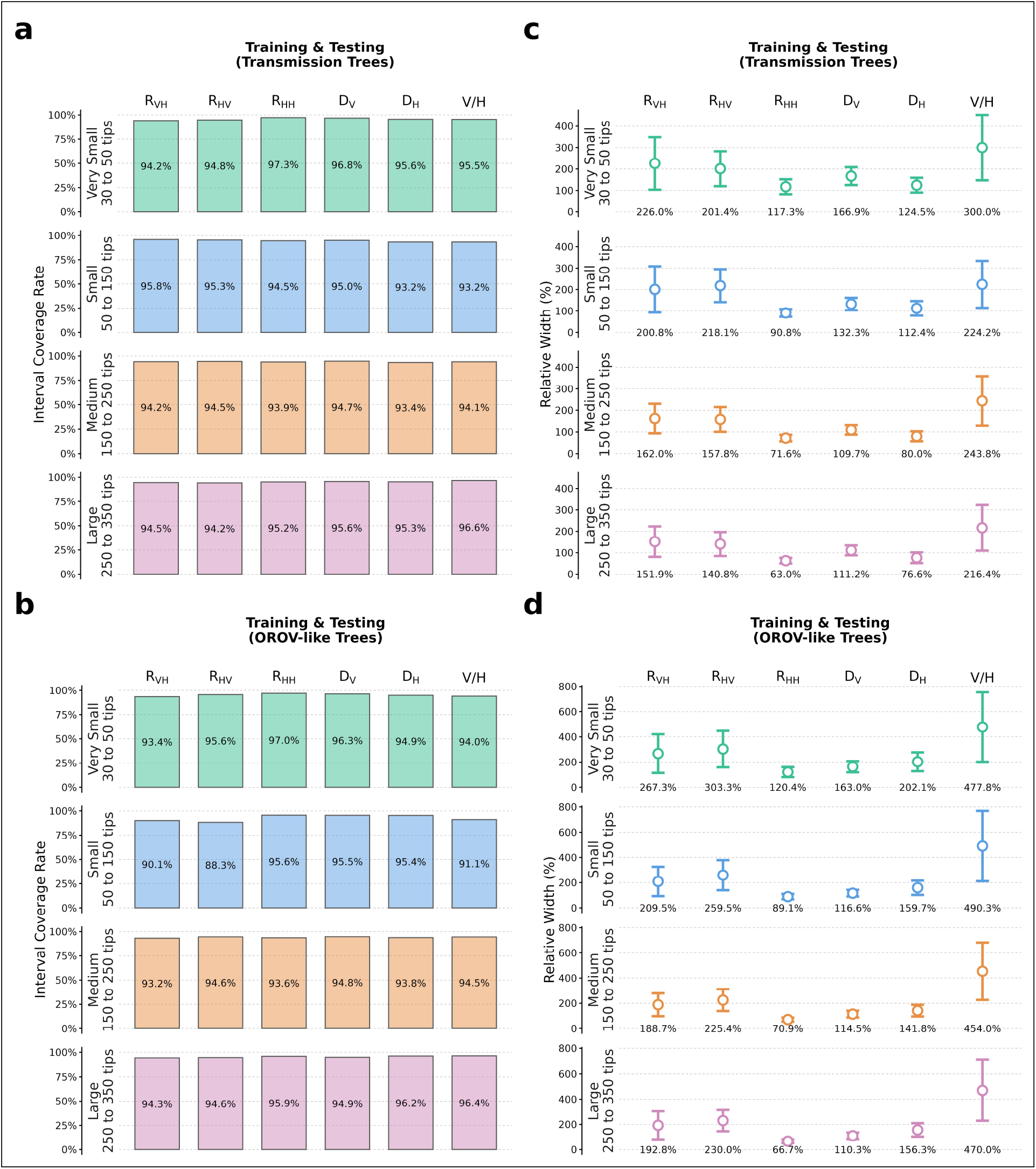
Coverage and relative width of 95% prediction intervals obtained with the deep learning implementation of the Birth–Death Exposed–Infectious Vector–Human (BDEIVH) model. **a**, b. Coverage of the 95% prediction intervals, defined as the proportion of true parameter values contained within the estimated intervals. c, d. Relative interval width, calculated as 100% × (*Q*_0.975_ − *Q*_0.025_)/(*true value*). Results are shown for the vector-to-human (*R_VH_*) and human-to-vector (*R_HV_*) reproduction numbers, the composite human-to-human reproduction number (*R_HH_*), the duration of infection in vectors (*D_V_*) and humans (*D_H_*), and the vector-to-human ratio (*V*/*H*), across four tree-size categories and for perfectly bifurcating trees (a, b) and OROV-like trees (c, d), in which internal branches were collapsed to reproduce reduced phylogenetic resolution.

## Supplementary Tables

**Table S1:**
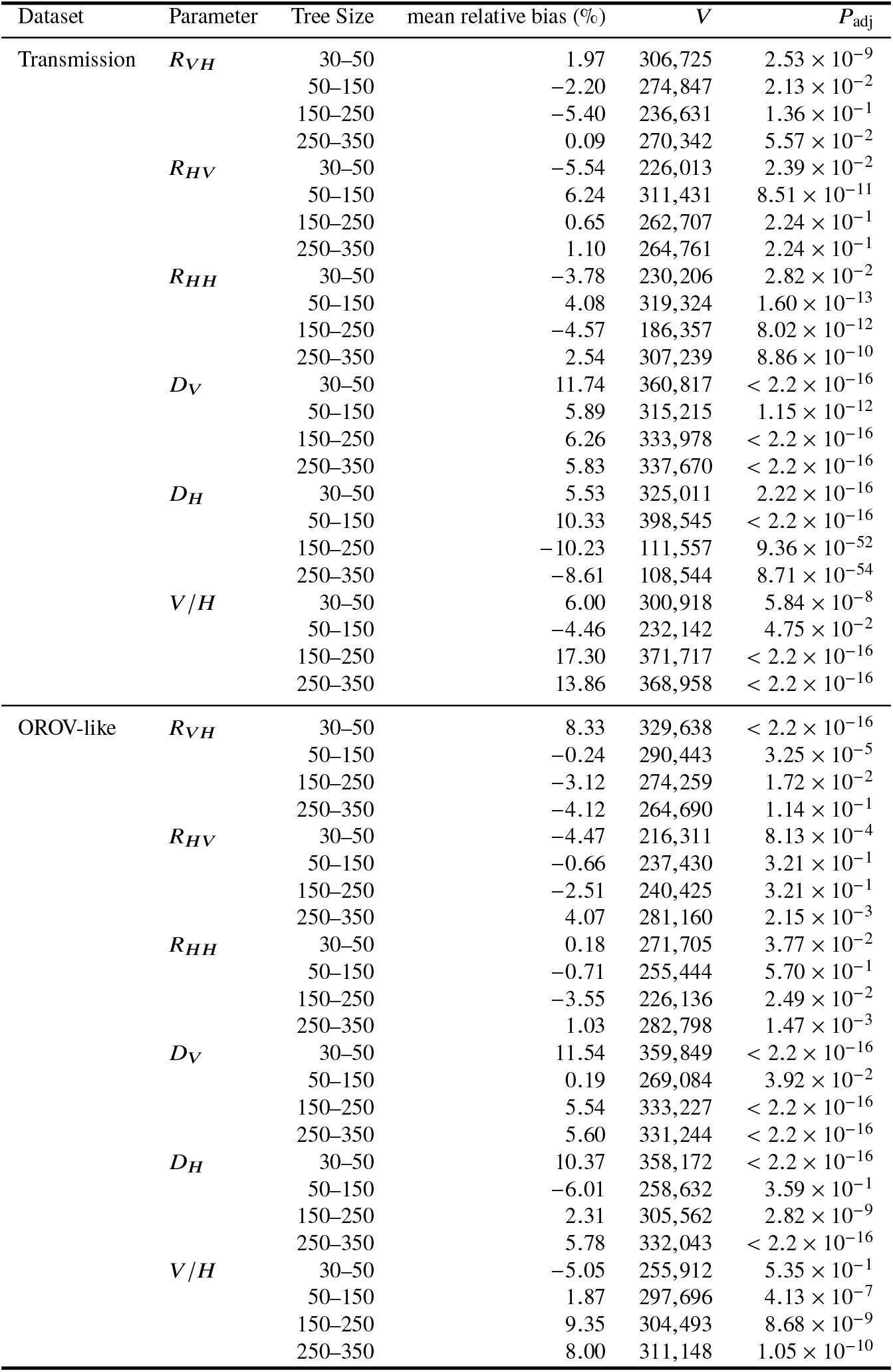
Wilcoxon signed-rank test results for relative bias across the four tree-size categories in the DL-implemented BDEIVH estimator.

| Dataset | Parameter | Tree Size | mean relative bias (%) | *V* | *P*<sub>adj</sub> |
|---|---|---|---|---|---|
| Transmission | *R<sub>VH</sub>* | 30–50 | 1.97 | 306,725 | 2.53 × 10<sup>−9</sup> |
|  |  | 50–150 | −2.20 | 274,847 | 2.13 × 10<sup>−2</sup> |
|  |  | 150–250 | −5.40 | 236,631 | 1.36 × 10<sup>−1</sup> |
|  |  | 250–350 | 0.09 | 270,342 | 5.57 × 10<sup>−2</sup> |
|  | *R<sub>HV</sub>* | 30–50 | −5.54 | 226,013 | 2.39 × 10<sup>−2</sup> |
|  |  | 50–150 | 6.24 | 311,431 | 8.51 × 10<sup>−11</sup> |
|  |  | 150–250 | 0.65 | 262,707 | 2.24 × 10<sup>−1</sup> |
|  |  | 250–350 | 1.10 | 264,761 | 2.24 × 10<sup>−1</sup> |
|  | *R<sub>HH</sub>* | 30–50 | −3.78 | 230,206 | 2.82 × 10<sup>−2</sup> |
|  |  | 50–150 | 4.08 | 319,324 | 1.60 × 10<sup>−13</sup> |
|  |  | 150–250 | −4.57 | 186,357 | 8.02 × 10<sup>−12</sup> |
|  |  | 250–350 | 2.54 | 307,239 | 8.86 × 10<sup>−10</sup> |
|  | *D<sub>V</sub>* | 30–50 | 11.74 | 360,817 | < 2.2 × 10<sup>−16</sup> |
|  |  | 50–150 | 5.89 | 315,215 | 1.15 × 10<sup>−12</sup> |
|  |  | 150–250 | 6.26 | 333,978 | < 2.2 × 10<sup>−16</sup> |
|  |  | 250–350 | 5.83 | 337,670 | < 2.2 × 10<sup>−16</sup> |
|  | *D<sub>H</sub>* | 30–50 | 5.53 | 325,011 | 2.22 × 10<sup>−16</sup> |
|  |  | 50–150 | 10.33 | 398,545 | < 2.2 × 10<sup>−16</sup> |
|  |  | 150–250 | −10.23 | 111,557 | 9.36 × 10<sup>−52</sup> |
|  |  | 250–350 | −8.61 | 108,544 | 8.71 × 10<sup>−54</sup> |
|  | *V*/*H* | 30–50 | 6.00 | 300,918 | 5.84 × 10<sup>−8</sup> |
|  |  | 50–150 | −4.46 | 232,142 | 4.75 × 10<sup>−2</sup> |
|  |  | 150–250 | 17.30 | 371,717 | < 2.2 × 10<sup>−16</sup> |
|  |  | 250–350 | 13.86 | 368,958 | < 2.2 × 10<sup>−16</sup> |
| OROV-like | *R<sub>VH</sub>* | 30–50 | 8.33 | 329,638 | < 2.2 × 10<sup>−16</sup> |
|  |  | 50–150 | −0.24 | 290,443 | 3.25 × 10<sup>−5</sup> |
|  |  | 150–250 | −3.12 | 274,259 | 1.72 × 10<sup>−2</sup> |
|  |  | 250–350 | −4.12 | 264,690 | 1.14 × 10<sup>−1</sup> |
|  | *R<sub>HV</sub>* | 30–50 | −4.47 | 216,311 | 8.13 × 10<sup>−4</sup> |
|  |  | 50–150 | −0.66 | 237,430 | 3.21 × 10<sup>−1</sup> |
|  |  | 150–250 | −2.51 | 240,425 | 3.21 × 10<sup>−1</sup> |
|  |  | 250–350 | 4.07 | 281,160 | 2.15 × 10<sup>−3</sup> |
|  | *R<sub>HH</sub>* | 30–50 | 0.18 | 271,705 | 3.77 × 10<sup>−2</sup> |
|  |  | 50–150 | −0.71 | 255,444 | 5.70 × 10<sup>−1</sup> |
|  |  | 150–250 | −3.55 | 226,136 | 2.49 × 10<sup>−2</sup> |
|  |  | 250–350 | 1.03 | 282,798 | 1.47 × 10<sup>−3</sup> |
|  | *D<sub>V</sub>* | 30–50 | 11.54 | 359,849 | < 2.2 × 10<sup>−16</sup> |
|  |  | 50–150 | 0.19 | 269,084 | 3.92 × 10<sup>−2</sup> |
|  |  | 150–250 | 5.54 | 333,227 | < 2.2 × 10<sup>−16</sup> |
|  |  | 250–350 | 5.60 | 331,244 | < 2.2 × 10<sup>−16</sup> |
|  | *D<sub>H</sub>* | 30–50 | 10.37 | 358,172 | < 2.2 × 10<sup>−16</sup> |
|  |  | 50–150 | −6.01 | 258,632 | 3.59 × 10<sup>−1</sup> |
|  |  | 150–250 | 2.31 | 305,562 | 2.82 × 10<sup>−9</sup> |
|  |  | 250–350 | 5.78 | 332,043 | < 2.2 × 10<sup>−16</sup> |
|  | *V*/*H* | 30–50 | −5.05 | 255,912 | 5.35 × 10<sup>−1</sup> |
|  |  | 50–150 | 1.87 | 297,696 | 4.13 × 10<sup>−7</sup> |
|  |  | 150–250 | 9.35 | 304,493 | 8.68 × 10<sup>−9</sup> |
|  |  | 250–350 | 8.00 | 311,148 | 1.05 × 10<sup>−10</sup> |

**Table S2:** Jonckheere–Terpstra test results for trends in relative error across the four tree-size categories in the DL-implemented BDEIVH estimator.

| Dataset | Parameter | JT | *P*-value | Decreasing trend |
|---|---|---|---|---|
| Transmission | *R*<sub>*VH*</sub> | 2,566,494 | < 2.2 × 10<sup>−16</sup> | Yes |
|  | *R*<sub>*HV*</sub> | 2,636,701 | < 2.2 × 10<sup>−16</sup> | Yes |
|  | *R*<sub>*HH*</sub> | 2,474,703 | < 2.2 × 10<sup>−16</sup> | Yes |
|  | *D*<sub>*V*</sub> | 2,671,659 | 4.44 × 10<sup>−16</sup> | Yes |
|  | *D*<sub>*H*</sub> | 2,592,591 | < 2.2 × 10<sup>−16</sup> | Yes |
|  | *V*/*H* | 2,785,800 | 7.77 × 10<sup>−8</sup> | Yes |
| OROV-like | *R*<sub>*VH*</sub> | 2,656,804 | < 2.2 × 10<sup>−16</sup> | Yes |
|  | *R*<sub>*HV*</sub> | 2,739,891 | 9.43 × 10<sup>−11</sup> | Yes |
|  | *R*<sub>*HH*</sub> | 2,554,794 | < 2.2 × 10<sup>−16</sup> | Yes |
|  | *D*<sub>*V*</sub> | 2,739,421 | 8.75 × 10<sup>−11</sup> | Yes |
|  | *D*<sub>*H*</sub> | 2,939,368 | 6.88 × 10<sup>−2</sup> | No |
|  | *V*/*H* | 2,902,366 | 8.40 × 10<sup>−3</sup> | Yes |

**Table S3:**
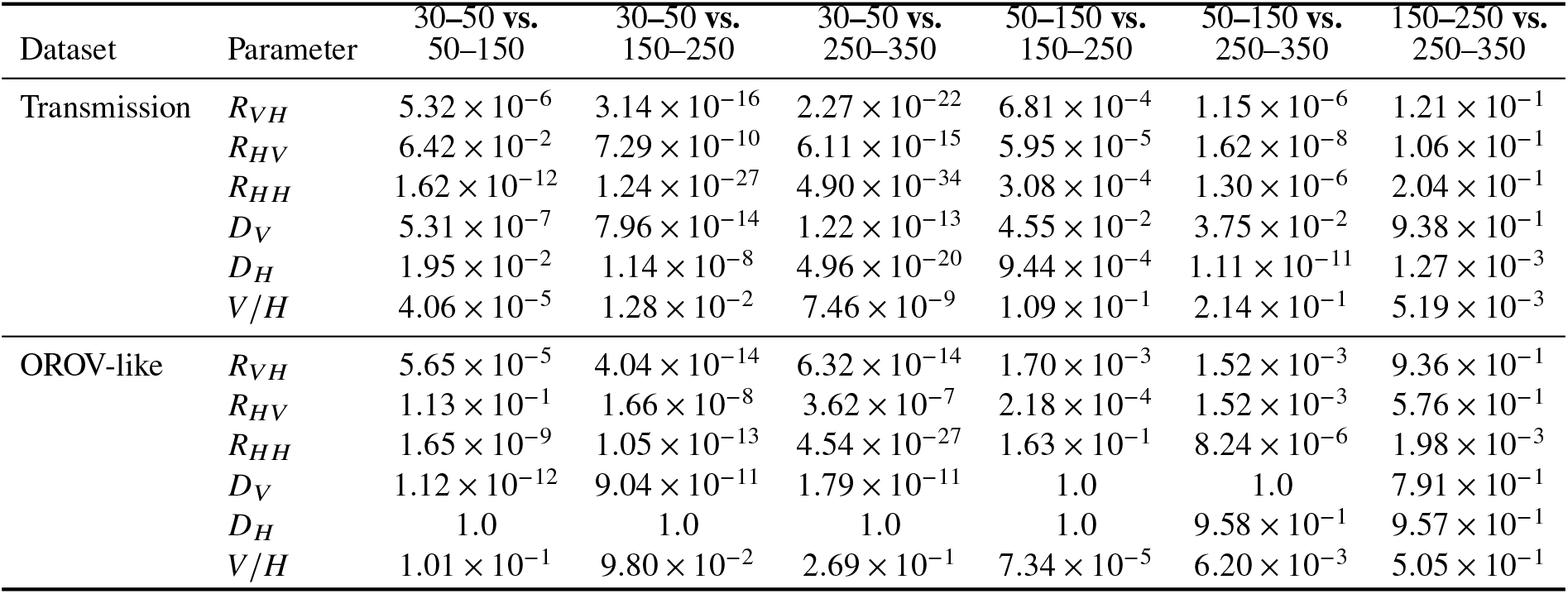
Dunn’s post-hoc test results for pairwise comparisons of relative error across the four tree-size categories in the DL-implemented BDEIVH estimator.

